# Predicting COVID-19 severity using major risk factors and received vaccines

**DOI:** 10.1101/2021.12.31.21268575

**Authors:** Ariel Israel, Alejandro A. Schäffer, Eugene Merzon, Ilan Green, Eli Magen, Avivit Golan-Cohen, Shlomo Vinker, Eytan Ruppin

**Affiliations:** Leumit Health Services, Israel; Cancer Data Science Laboratory, Center for Cancer Research, National Cancer Institute, Bethesda, MD USA; Adelson School of Medicine, Ariel University, Ariel, Israel; Department of Family Medicine, Sackler Faculty of Medicine, Tel-Aviv University, Israel; Medicine C Department, Clinical Immunology and Allergy Division, Barzilai University Medical Center, Ben-Gurion University of the Negev, Ashkelon, Israel

**Keywords:** COVID-19, disease severity, calculator, diabetes, obesity, kidney disease

## Abstract

**Background:** Vaccines are highly effective in preventing severe disease and death from COVID-19, and new medications that can reduce severity of disease have been approved. However, many countries are facing limited supply of vaccine doses and medications. A model estimating the probabilities for hospitalization and mortality according to individual risk factors and vaccine doses received could help prioritize vaccination and yet scarce medications to maximize lives saved and reduce the burden on hospitalization facilities.

**Methods:** Electronic health records from 101,039 individuals infected with SARS-CoV-2, since the beginning of the pandemic and until November 30, 2021 were extracted from a national healthcare organization in Israel. Logistic regression models were built to estimate the risk for subsequent hospitalization and death based on the number of BNT162b2 mRNA vaccine doses received and few major risk factors (age, sex, body mass index, hemoglobin A1C, kidney function, and presence of hypertension, pulmonary disease and malignancy).

**Results:** The models built predict the outcome of newly infected individuals with remarkable accuracy: area under the curve was 0.889 for predicting hospitalization, and 0.967 for predicting mortality. Even when a breakthrough infection occurs, having received three vaccination doses significantly reduces the risk of hospitalization by 66% (OR=0.339) and of death by 78% (OR=0.223).

**Conclusions:** The models enable rapid identification of individuals at high risk for hospitalization and death when infected. These patients can be prioritized to receive booster vaccination and the yet scarce medications. A calculator based on these models is made publicly available on http://covidest.web.app

## Introduction

Since the start of the COVID-19 pandemic, over 250 million individuals have been infected and over 5 million individuals have died (https://coronavirus.jhu.edu/map.html). Since late 2020, multiple vaccines have been developed^1,2^, and more recently, various treatments including monoclonal antibodies, and anti-viral molecules have been FDA approved^3^. Unfortunately, there are still not enough vaccine doses and medications available worldwide. Thus, it is an urgent unmet need to identify objectively which patients would most benefit from booster vaccination doses or early access to the yet scarce medicines^4–6^.

To address these questions, we conducted a retrospective study among members of Leumit Health Services (LHS), one of the four main health maintenance organizations (HMOs) in Israel, which has over 700,000 members. Our study starts at the beginning of the pandemic and hence covers periods before any vaccines were available and after. Israel was one of the first countries to implement a large-scale vaccination plan (using Pfizer/BioNTech BNT162b2 vaccine)^7^ and to deploy a third vaccine booster dose. Nevertheless, there exists a large variation in vaccination uptake^8^, which provides an opportunity to assess the beneficial effects of vaccination in conjunction with other patient risk factors. In addition to vaccinations, we considered other traits associated with COVID-19 severity, including type II diabetes ^9–14^, kidney disease ^11,14–16^, chronic obstructive pulmonary disease (COPD) ^16–20^, obesity^10–12,21,22^, hypertension ^23–25^, malignancy^26^ and advanced age ^11,27–29^.

We constructed predictive models that estimate the risks that patients newly infected with SARS-CoV-2 (as reflected by positive PCR tests) would require hospitalization during the disease course and would die from COVID-19. Predictions are based on patient’s age, sex, the clinical factors mentioned above and vaccination status at the time of infection (0,1,2, or 3 doses). Importantly, all these factors are typically part of a patient’s medical records and measured in routine laboratory testing.

Models were built based on the outcomes of LHS COVID-19 patients during the study period. To keep the models simple and interpretable, and to allow deployment of the models in any health provider information system, we used multivariable logistic regression models based on the most essential risk factors. Regression coefficients and odds ratio (OR) for each factor are provided, and these can be calculated using a simple formula to obtain risk estimates for any individual. The resulting models additionally allow to estimate the decrease in hospitalization and mortality risk that can be achieved for any individual by providing additional vaccine doses. These estimates can help identify individuals and segments of population for which additional vaccination could help most. Notably, most treatments available should be given early in the course of the disease to those at high risk, to be most effective. A web-based calculator is provided, and the approach to run or adapt the models is fully described.

## Methods

### Study subjects and study design

This is a population study performed in Leumit Health Services (LHS), a nationwide healthcare provider in Israel, which provides services to around 700,000 members throughout the country. LHS uses centrally managed electronic health records (EHRs), continuously updated regarding subjects’ demographics, medical diagnoses, medical encounters, hospitalizations and laboratory tests. All LHS members have similar health insurance and similar access to healthcare services.

The study is based on members of LHS, of age ≥ 5 (eligible for vaccination), who had at least one positive PCR test for SARS-CoV-2 between April 2020 and November 30, 2021. Patients’ data were extracted from LHS central data warehouse on January 3, 2022. For each COVID-19 episode, the date of the first positive PCR test was taken as the index date. Number of vaccine doses received were calculated at the index date. Diagnosis and laboratory data were queried as known 15 days before the index date. The following factors were included in the analysis: sex, age, the Body Mass Index (BMI) as a categorical variable (<18.5; 18.5-25; 25-30; 30-35; ≥35), hemoglobin A1C range (<6.5; 6.5-8; 8-10; ≥10), last glomerular filtration rate (GFR) as an estimate of kidney function as a categorical variable (categories: ≥90; 60-89; 45-59; 30-44; <30). Presence of comorbidity conditions was assessed by presence of an active chronic diagnosis at this date. Chronic diagnoses, coded according to the International Classification of Diseases 9th revision (ICD-9), are regularly added, updated or ended, by the treating physician, at each encounter. The validity of chronic diagnoses in the registry has been previously examined and confirmed as high^30–32^.

To keep the models as simple as possible, we deliberately limited the models to the conditions that we identified as having the most significant effect on disease severity: hypertension, chronic obstructive pulmonary disease (COPD) and malignancy (solid or hematologic). Individuals who had a pregnancy diagnosis up to 210 days before the PCR test were excluded, as hospitalization would often pertain to pregnancy surveillance or delivery, and not reflect disease severity.

### Ethics statement

The study protocol was approved by the statutory clinical research committee of Leumit Health Services and the Shamir Medical Center Institutional Review Board (129-2-LEU). Informed consent was waived because this is a large-scale retrospective study and all data were deidentified.

### SARS-CoV-2 testing by real-time RT-PCR

Nasopharyngeal swabs were taken and examined for SARS-CoV-2 by real-time RT-PCR performed with internal positive and negative controls, according to World Health Organization guidelines, using TaqPath™ COVID-19 Combo Kit (Thermo Fisher Scientific) and COBAS SARS-Cov-2 6800/8800 (Roche Diagnostics) assays.

### Statistical analyses

Standard descriptive statistics were used to present the demographic characteristics of individuals included in the study cohort. Statistical analyses were done with R version 4.0.4 (R Foundation for Statistical Computing). Multivariable logistic regression models were fitted using the “glm” procedure, using age as a continuous variable, sex as a binary variable, number of vaccine doses, BMI category, hemoglobin A1C range, and GFR estimate^33^ as categorical variables; and presence of hypertension, pulmonary disease or malignancy as binary variables. Receiver operating characteristic (ROC) curves were used to assess model performance^34^, using 10-fold cross-validation. A two-sided P<0.05 was considered statistically significant. Missing values, which appeared only in BMI, kidney function and hemoglobin A1C variables, were treated by two different approaches. We used k-nearest-neighbors imputation to replace missing values, and also performed an alternative analysis in which missing values were treated as separate “missing” categories. We display here the regression coefficients obtained after imputation. Both methods resulted in very similar classifier performance.

## Results

### Factors associated with hospitalization of SARS-CoV-2 positive individuals

101,039 COVID-19 episodes were included, based on a positive test for SARS-CoV-2 obtained between April 1, 2020 and November 30, 2021. 393 (0.4%) resulted in patient death during hospitalization or within 30 days of the disease, and 1,752 (1.7%) required patient hospitalization for COVID-19 that did not end in patient death.

**Table 1A** shows the baseline characteristics of individuals included in the cohort, according to their outcomes. **Table 1B** displays the distribution of categorical variables after imputation of missing values. Generally, patients hospitalized and who died of the disease were older, had a greater proportion of males, had higher BMIs and hemoglobin A1C values, and were more likely to be affected with hypertension, pulmonary disease, malignancy, and impaired kidney function.

**Table 1A:** Demographic and clinical characteristics of the study population.

|  |  | **Not hospitalized** | **Hospitalized**<br>**(not deceased)** | **Deceased** |
| --- | --- | --- | --- | --- |
| **N (%)** |  | 98,894 (97.9%) | 1,752 (1.7%) | 393 (0.4%) |
| **Vaccines doses** | **0** | 82,261 (83.2 %) | 1,405 (80.2 %) | 295 (75.1 %) |
|  | **1** | 4,732 (4.8 %) | 138 (7.9 %) | 32 (8.1 %) |
|  | **2** | 10,436 (10.6 %) | 176 (10.0 %) | 61 (15.5 %) |
|  | **3** | 1,465 (1.5 %) | 33 (1.9 %) | 5 (1.3 %) |
| **Sex** | **Female** | 48,565 (49.1 %) | 798 (45.5 %) | 169 (43.0 %) |
| **Age** | **mean (SD)** | 29.44 (19.17) | 58.44 (19.03) | 75.27 (13.06) |
| **Age category** | **5-11** | 19,603 (19.8 %) | 19 (1.1 %) | 0 (0.0 %) |
|  | **12-17** | 15,999 (16.2 %) | 30 (1.7 %) | 0 (0.0 %) |
|  | **18-39** | 34,374 (34.8 %) | 245 (14.0 %) | 6 (1.5 %) |
|  | **40-59** | 20,361 (20.6 %) | 561 (32.0 %) | 44 (11.2 %) |
|  | **≥ 60** | 8,557 (8.7 %) | 897 (51.2 %) | 343 (87.3 %) |
| **Comorbidities** | **Hypertension** | 9,321 (9.4 %) | 880 (50.2 %) | 301 (76.6 %) |
|  | **Pulmonary Disease** | 1,592 (1.6 %) | 167 (9.5 %) | 74 (18.8 %) |
|  | **Malignancy** | 2,258 (2.3 %) | 202 (11.5 %) | 83 (21.1 %) |
| **BMI category** | **Underweight < 18.5** | 22,506 (22.8 %) | 44 (2.5 %) | 10 (2.5 %) |
|  | **Normal 18.5 - 25** | 32,373 (32.7 %) | 283 (16.2 %) | 83 (21.1 %) |
|  | **Overweight 25 - 30** | 21,396 (21.6 %) | 566 (32.3 %) | 126 (32.1 %) |
|  | **Obese I 30 - 35** | 10,763 (10.9 %) | 444 (25.3 %) | 85 (21.6 %) |
|  | **Obese II+ ≥ 35** | 5,493 (5.6 %) | 372 (21.2 %) | 72 (18.3 %) |
|  | **\*missing\*** | 6,363 (6.4 %) | 43 (2.5 %) | 17 (4.3 %) |
| **Kidney function**<br>**GFR category** | **G1 (Normal) ≥ 90** | 64,097 (64.8 %) | 850 (48.5 %) | 95 (24.2 %) |
|  | **G2 60-89** | 12,622 (12.8 %) | 596 (34.0 %) | 142 (36.1 %) |
|  | **G3a 45-59** | 840 (0.8 %) | 140 (8.0 %) | 68 (17.3 %) |
|  | **G3b 30-44** | 263 (0.3 %) | 74 (4.2 %) | 46 (11.7 %) |
|  | **G4/G5 <30** | 151 (0.2 %) | 45 (2.6 %) | 37 (9.4 %) |
|  | **\*missing\*** | 20,921 (21.2 %) | 47 (2.7 %) | 5 (1.3 %) |
| **Hemoglobin A1C**<br>**range** | **< 6.5** | 38,743 (92.2 %) | 1,106 (72.2 %) | 268 (70.7 %) |
|  | **[6.5, 8.0)** | 2,129 (5.1 %) | 253 (16.5 %) | 70 (18.5 %) |
|  | **[8.0, 10.0)** | 815 (1.9 %) | 115 (7.5 %) | 31 (8.2 %) |
|  | **≥ 10.0** | 328 (0.8 %) | 58 (3.8 %) | 10 (2.6 %) |
|  | **\*missing\*** | 56,879 (57.5 %) | 220 (12.6 %) | 14 (3.6 %) |

**Table 1B:** Clinical characteristics of the study population after missing variables imputation.

|  |  |  |  |  |  |
| --- | --- | --- | --- | --- | --- |
| **BMI category** | **Underweight** | **< 18.5** | 25,090 (25.4 %) | 47 (2.7 %) | 10 (2.5 %) |
|  | **Normal** | **18.5 - 25** | 34,615 (35.0 %) | 294 (16.8 %) | 85 (21.6 %) |
|  | **Overweight** | **25 - 30** | 22,687 (22.9 %) | 593 (33.8 %) | 139 (35.4 %) |
|  | **Obese I** | **30 - 35** | 11,006 (11.1 %) | 446 (25.5 %) | 87 (22.1 %) |
|  | **Obese II+** | **≥ 35** | 5,496 (5.6 %) | 372 (21.2 %) | 72 (18.3 %) |
| **Kidney function**<br>**GFR category** | **G1 (Normal)** | **≥ 90** | 84,503 (85.4 %) | 887 (50.6 %) | 90 (22.9 %) |
|  | **G2** | **60-89** | 13,124 (13.3 %) | 590 (33.7 %) | 144 (36.6 %) |
|  | **G3a** | **45-59** | 860 (0.9 %) | 141 (8.0 %) | 71 (18.1 %) |
|  | **G3b** | **30-44** | 256 (0.3 %) | 87 (5.0 %) | 47 (12.0 %) |
|  | **G4/G5** | **<30** | 151 (0.2 %) | 47 (2.7 %) | 41 (10.4 %) |
| **Hemoglobin A1C**<br>**range** |  | **< 6.5** | 95,481 (96.5 %) | 1,322 (75.5 %) | 282 (71.8 %) |
|  |  | **[6.5, 8.0)** | 2,266 (2.3 %) | 257 (14.7 %) | 70 (17.8 %) |
|  |  | **[8.0, 10.0)** | 819 (0.8 %) | 115 (6.6 %) | 31 (7.9 %) |
|  |  | **≥ 10.0** | 328 (0.3 %) | 58 (3.3 %) | 10 (2.5 %) |

We built multivariable logistic regression models to predict both the hospitalization and mortality outcomes. The odds ratios from multivariable regression models reflect the extent with which each risk factor affects the outcome, after adjustment for the others.

**Table 2** displays the model for hospitalization risk based on the comparison of 2,145 episodes that resulted either in hospitalization or death vs. 98,894 infections that did not necessitate hospitalization. For each variable, the regression coefficient, with the corresponding odds ratio and 95% confidence interval and p-values are displayed. The footnote explains how to calculate the outcome probability for any given patient data.

**Table 2:**
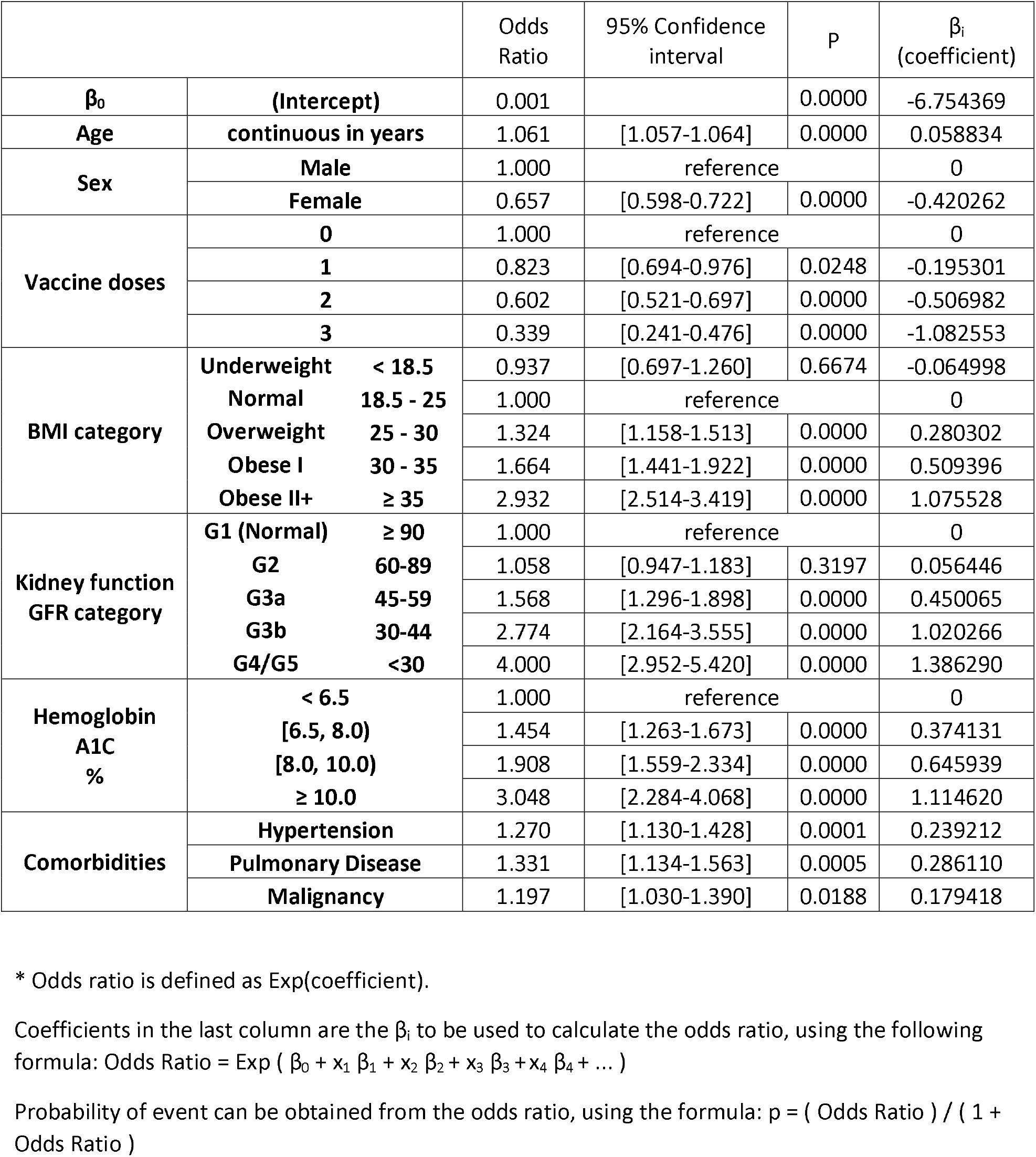
Logistic regression model for hospitalization risk.

We emphasize a few key findings arising from the multivariable regression analysis. First, increased age is significantly associated with the risk of hospitalization: each year of age increases the odds for hospitalization by a multiplicative factor of 1.061, which means that compared to an individual aged twenty with similar other risk factors, a patient aged sixty is 10 times more likely to get hospitalized, and a patient aged eighty, 34 times more likely to get hospitalized. Female sex reduces odds for hospitalization by 34%. Obesity increases its risk in a gradual manner (OR=1.324 for BMI between 25 and 30, OR=1.664 for BMI between 30-35, and OR=2.932 for BMI over 35, compared to the reference category of normal BMI, p<0.001 for all). Diabetes mellitus, as reflected by most recent hemoglobin A1C values is independently associated with increased risk in a gradual manner (OR=1.454 for A1C between 6.5 and 8%, OR=1.908 for A1C between 8 and 10%, and OR=3.048 for A1C above 10% compared to the reference category of A1C below 6.5%, p<0.001 for all). Impaired kidney function is also associated with increased risk in a gradual manner (OR=1.568 for GFR between 45 and 59, OR=2.774 for GFR between 30 and 44, and OR=4.000 for GFR below 30 compared to the reference category of GFR above 90, p<0.001 for all). Hospitalization risks significantly increase with hypertension (OR=1.270, 95% CI [1.130-1.428]), pulmonary disease (OR=1.331, 95% CI [1.134-1.563]) and cancer (OR=1.197, 95% CI [1.030-1.390]).

As expected, compared to unvaccinated patients, vaccination significantly decreases the hospitalization risk with (OR= 0.602, 95% CI [0.521-0.697] for two vaccine doses and OR=0.339, 95% CI [0.241-0.476] for three vaccine doses, p<0.001 for both categories), and even for the relatively small group of single vaccinated individuals, with OR=0.823, 95% CI [0.694-0.976] (P=0.025).

The ROC curve shows the diagnostic ability of a classifier as its discrimination threshold is varied. We performed a 10-fold cross validation and calculated the ROC curve to estimate the performance of our model (**Figure 1**). The performance of the hospitalization risk model was remarkably accurate, with an area under the curve (AUC) of 0.889. The model was able to predict 50%, 80% and 90% of hospitalizations, with respective specificities of 95.3%, 82.2% and 70.2%.

**Figure 1:**
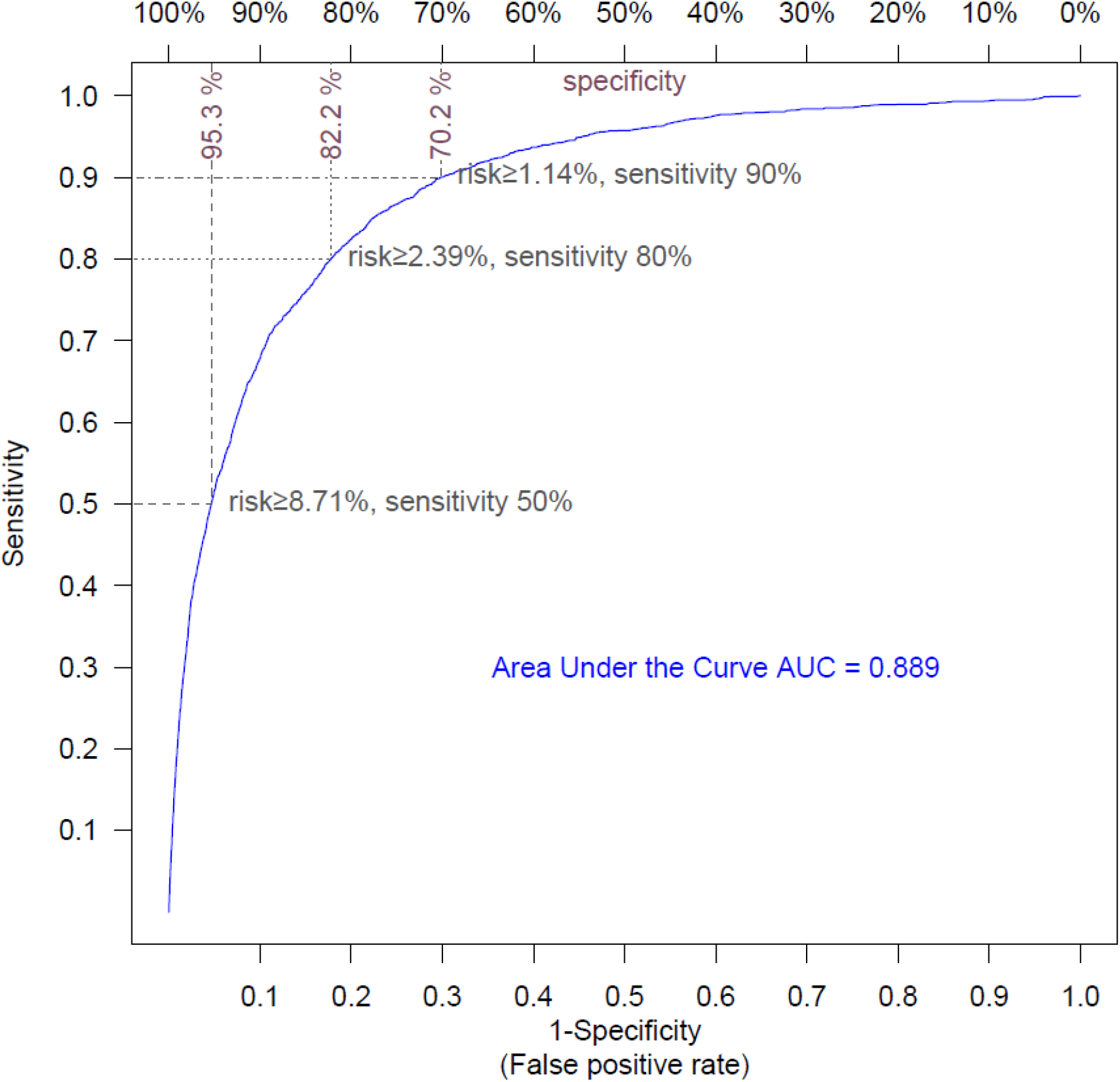
Receiver Operating Curve for hospitalization risk model. The ROC curve shows the sensitivity and the specificity of the hospitalization model as its discrimination threshold is varied. With a threshold of 8.71% for risk, 50% of the COVID-19 episodes necessitating hospitalization can be identified (sensitivity=50%), and specificity is 95.3% (false positive rate=4.7%); with a risk threshold of 2.39%, sensitivity is 80% and specificity is 82.2% (false positive rate=7.8%); with a risk threshold of 1.14%, sensitivity is 90% and specificity is 70.2% (false positive rate=30.8%).

### Factors associated with mortality for SARS-CoV-2 positive individuals

**Table 3** displays the model for mortality risk. It is based on the comparison of 393 fatal cases of COVID-19 compared to 101,039 disease episodes that did not end in patient death. The smaller size of the outcome group limits the power of the model, nevertheless a few factors are associated with a large statistically significant effect: Advanced age is even more strikingly associated with increased mortality risk. Each year of age increases the odds for death by a factor of 1.105: compared to an individual aged twenty with similar other risk factors, a patient aged sixty is 54 times more likely to die of the disease, and a patient aged eighty is 393 times more likely to die. Female sex is also associated with reduced risk of death, reducing this risk by about a half.

**Table 3:** Logistic regression model for mortality risk.

| | | Odds Ratio | 95% Confidence interval | P | $\beta_i$<br>(coefficient) |
| --- | --- | --- | --- | --- | --- |
| $\beta_0$ | (Intercept) | 0.000 | | 0.0000 | -11.227376 |
| Age | continuous in years | 1.105 | [1.095-1.115] | 0.0000 | 0.099573 |
| Sex | Male | 1.000 | reference |  | 0 |
|  | Female | 0.500 | [0.401-0.625] | 0.0000 | -0.692446 |
| Vaccine doses | 0 | 1.000 | reference |  | 0 |
|  | 1 | 0.921 | [0.627-1.354] | 0.6771 | -0.081842 |
|  | 2 | 0.936 | [0.698-1.254] | 0.6561 | -0.066541 |
|  | 3 | 0.223 | [0.091-0.551] | 0.0011 | -1.498783 |
| BMI category | Underweight < 18.5 | 2.179 | [1.056-4.496] | 0.0350 | 0.778997 |
|  | Normal 18.5 - 25 | 1.000 | reference |  | 0 |
|  | Overweight 25 - 30 | 0.979 | [0.733-1.307] | 0.8866 | -0.021027 |
|  | Obese I 30 - 35 | 1.085 | [0.785-1.500] | 0.6196 | 0.081961 |
| | Obese II+ $\geq 35$ | 1.963 | [1.383-2.786] | 0.0002 | 0.674479 |
| Kidney function<br>GFR category | G1 (Normal) $\geq 90$ | 1.000 | reference | | 0 |
|  | G2 60-89 | 1.283 | [0.965-1.705] | 0.0861 | 0.249162 |
|  | G3a 45-59 | 2.000 | [1.390-2.878] | 0.0002 | 0.693180 |
|  | G3b 30-44 | 3.097 | [2.035-4.715] | 0.0000 | 1.130578 |
|  | G4/G5 <30 | 6.888 | [4.389-10.810] | 0.0000 | 1.929831 |
| Hemoglobin<br>A1C<br>% | < 6.5 | 1.000 | reference |  | 0 |
|  | [6.5, 8.0) | 1.137 | [0.851-1.518] | 0.3842 | 0.128408 |
|  | [8.0, 10.0) | 1.479 | [0.983-2.226] | 0.0602 | 0.391618 |
| | $\geq 10.0$ | 1.782 | [0.905-3.510] | 0.0948 | 0.577767 |
| Comorbidities | Hypertension | 1.348 | [1.011-1.797] | 0.0421 | 0.298497 |
|  | Pulmonary Disease | 1.475 | [1.113-1.956] | 0.0069 | 0.388824 |
|  | Malignancy | 1.138 | [0.868-1.491] | 0.3489 | 0.129199 |

\* Odds ratio is defined as $\text{Exp}(\text{coefficient})$ .
Coefficients in the last column are the $\beta_i$ to be used to calculate the odds ratio, using the following formula: $\text{Odds Ratio} = \text{Exp} ( \beta_0 + x_1 \beta_1 + x_2 \beta_2 + x_3 \beta_3 + x_4 \beta_4 + \dots )$
Probability of event can be obtained from the odds ratio, using the formula: $p = ( \text{Odds Ratio} ) / ( 1 + \text{Odds Ratio} )$

Both extreme obesity and underweight increased the risk of death (OR=1.963 for BMI above 35, and OR=2.179 for BMI below 18.5, compared to normal BMI), while other BMI categories were not significantly associated with death. Impaired kidney function was also associated with augmented mortality risk in a gradual manner (OR=2.000 for GFR between 45 and 59, OR=3.097 for GFR between 30 and 44, and OR=6.888 for GFR below 30 compared to the reference category of GFR above 90, p<0.001 for all). Diabetes mellitus, as reflected by last hemoglobin A1C also increases mortality risk in a gradual manner, although more moderately than for hospitalization. The other comorbidities coefficients are overall similar to those for hospitalization, although the smaller outcome group allowed to detect statistical significance only for hypertension and pulmonary disease. Vaccination with a booster dose significantly decreased the mortality risk by 78% (OR= 0.223, 95% CI [0.091-0.551], p=0.001).

We performed a 10-fold cross validation and plotted the ROC curve to estimate the performance of the mortality risk model (**Figure 2**). The model was very accurate, with an AUC of 0.967, and was able to predict 50%, 80% and 90% of death events, with respective specificities of 98.6%, 95.2% and 91.2%.

**Figure 2:**
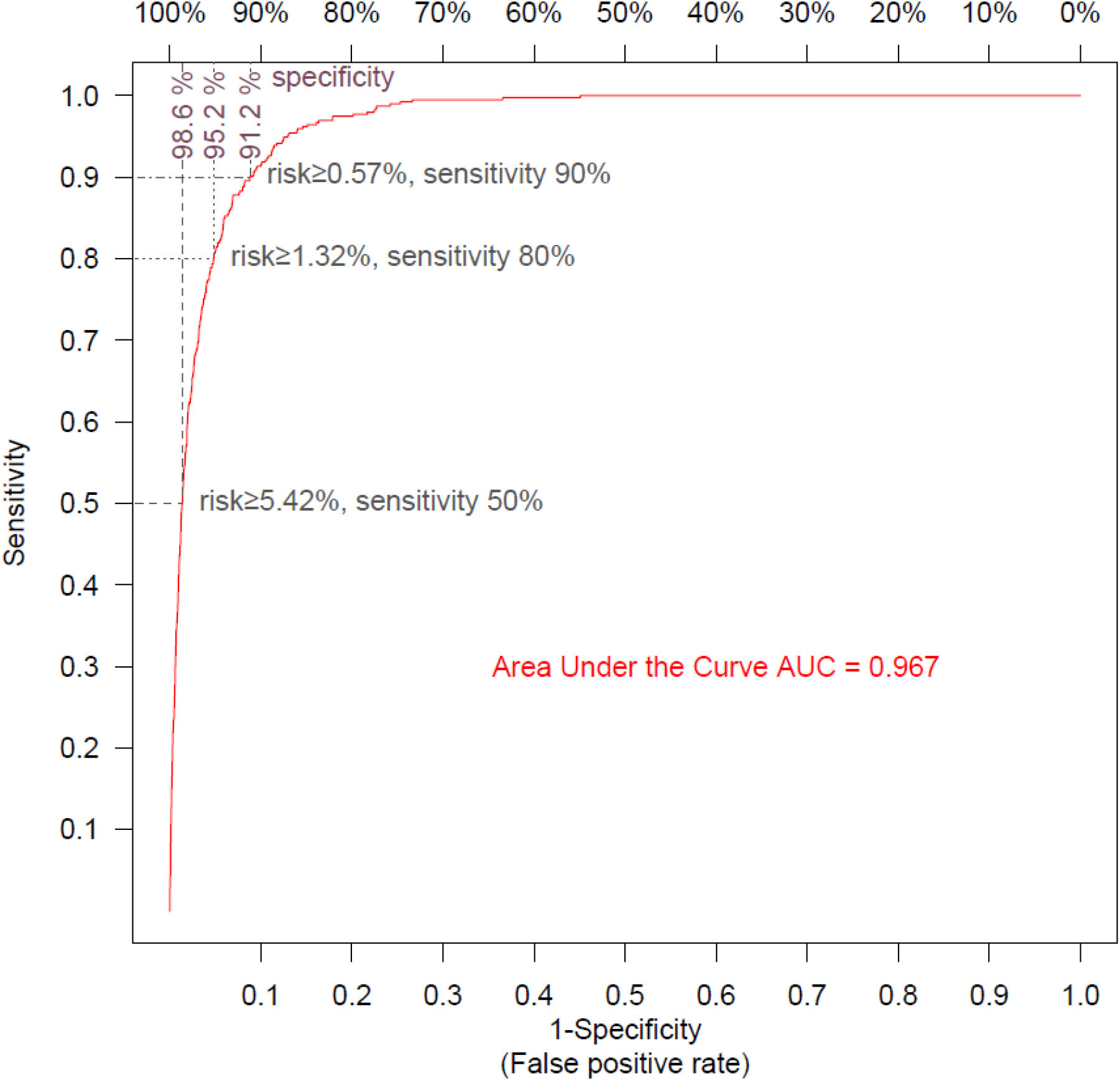
Receiver Operating Curve for mortality risk model. The ROC curve shows the sensitivity and the specificity of the mortality model as its discrimination threshold is varied. With a threshold of 5.42% for risk, 50% of the COVID-19 episodes ending in patient death can be identified (sensitivity=50%), and specificity is 98.6% (false positive rate=1.4%); with a risk threshold of 1.32%, sensitivity is 80% and specificity is 95.2% (false positive rate =4.8%); with a risk threshold of 0.57%, sensitivity is 90% and specificity is 91.2% (false positive rate=8.8%).

### Risk calculators

**Tables 2 and 3** provide all the coefficients, as well as the formula that is used to calculate the absolute risk of a given individual, using the regression models described above. Basically, the coefficients are to be multiplied by the corresponding variables and summed to obtain the natural logarithm of the odds ratio. After exponentiation, the odds ratio can be converted to a probability by dividing it by its value plus one. This calculator is made available online and can be used to calculate hospitalization and mortality probabilities for any given individual.

## Discussion

This study developed models that can estimate the risks of subsequent hospitalization and death for any individual newly infected with SARS-CoV-2, using health records from a large healthcare provider in Israel. It also provides further large-scale confirmation of recently published studies from other Israeli health care providers that showed that a third, booster vaccine provides a sharp and almost immediate increase in protection^35,36^. Considering that vaccination has also been shown to substantially decrease the risk for symptomatic infection, its overall cumulative effect on hospitalization and death are even greater than the odds ratio reported here.

Our study capitalizes on the centrally managed, comprehensive electronic health records maintained in our health organization, that are continuously updated with hospitalization and death from COVID-19; the early adoption of a homogenous vaccination program in Israel, and of the booster dose; and the relatively large number of individuals that have been tested positive for COVID-19. We deliberately opted for model simplicity by limiting the number of input variables to recognized clinical factors associated with disease severity that are documented in most health organizations. We intentionally did not include country specific demographic variables, to generate a model that is generalizable to other populations. Even with these limited inputs, the resulting AUCs are highly accurate, achieving 0.889 for hospitalization and 0.967 for mortality. The better AUC performance of the model for mortality risk likely reflects that death is mostly determined by the patient health status, while the decision to hospitalize a patient is additionally impacted by availability of family or social support at home and the patient’s own preferences, which are not accounted for by our model. Importantly, these mortality and hospitalization risk estimates can be given as soon as the disease is diagnosed, allowing to identify which patients are most at risk for a severe course, so that they could be given treatment early in the infection. The models can also prioritize which populations to vaccinate, or urged to receive booster doses, to maximize lives saved and to reduce the load on hospitalization facilities. Several attempts have already been made to build predictive models for COVID-19 severity, notably ^4,16,27,37,38^. Nevertheless, there is still an unmet need for simple models, which like CURB-65^39^, could allow instant triage of new patients, using few essential parameters, including vaccination status. This approach allowed us to produce models with remarkably high accuracy.

Importantly, in contrast to what we and others found for the infection risk^30^, we did not observe that time elapsed since vaccination significantly increased hospitalization and mortality risks. For these outcomes, the protective effect of vaccination was largely determined by the cumulative number of vaccine doses received. This may indicate that the immune system response elicited by mRNA vaccine injection has a more lasting effect on hospitalization and mortality risk than on the risk of symptomatic infection following exposure to the virus.

Our study has several limitations. First, it is based on a population which was vaccinated almost exclusively with the Pfizer/Biotech BNT162b2 vaccine, and with the first two doses spaced by 21 days. It is uncertain how the estimated effect of vaccination under these conditions would apply to populations vaccinated using different vaccines or using a different vaccination schedule. Moreover, factors specific to our health organization may have affected the results, such as criteria for hospital admission, and treatment decisions that influence mortality. Evolving patient management policies could have a confounding effect on the number of vaccine doses at different times. Last, we have yet to assess the model ability to predict disease severity with new viral variants, such as the recently spreading Omicron. Additional studies in different populations would help to ascertain the validity of our models in different settings. To enable such validation studies, we provide the full model formulas and encourage their use.

In conclusion, the models described here, and available online as a calculator, allow to identify individuals most at risk for severe disease or death if infected, using very few essential parameters and vaccination status. This approach can guide public health decisions to optimally allocate vaccines and scarce medicines to maximize lives saved^5^.

Calculator address: https://covidest.web.app/

## Data Availability

Data were obtained from patients' electronic health records and IRB approval restrains its use to researchers inside Leumit Health Services.

https://covidest.web.app/calculator.html

## Abbreviations

AUC: Area Under the Curve
BMI: Body mass Index
COPD: chronic obstructive pulmonary disease
COVID-19: Coronavirus disease of 2019
GFR: glomerular filtration rate
LHS: Leumit Health Services
OR: odds ratio
ROC: Receiver Operating Characteristic
SARS-CoV-2: Severe acute respiratory syndrome coronavirus 2

## Acknowledgments

This research was supported in part by the Intramural Research Program, National Institutes of Health, National Cancer Institute, Center for Cancer Research. The content of this publication does not necessarily reflect the views or policies of the Department of Health and Human Services, nor does mention of trade names, commercial products, or organizations imply endorsement by the U.S. Government. Thanks to Dr. Doug Lowy for helpful comments.

The authors had no conflict of interest to report

## Notes

### Competing Interest Statement

The authors have declared no competing interest.

### Funding Statement

This research was supported in part by the Intramural Research Program of the National Institutes of Health, National Cancer Institute.

### Author Declarations

The Institutional Review Board (IRB) of Shamir Medical Center gave ethical approval for this work (129-2-LEU).

### Summary of Updates

Added k-nearest-neighbors imputation for missing data; Minor english corrections

